# Association of Physical Activity Volume and Intensity with Incident Cardiovascular Disease: a UK Biobank Study

**DOI:** 10.1101/2022.02.23.22271386

**Authors:** Paddy C. Dempsey, Alex V. Rowlands, Tessa Strain, Francesco Zaccardi, Nathan Dawkins, Cameron Razieh, Melanie J. Davies, Kamlesh K. Khunti, Charlotte L. Edwardson, Katrien Wijndaele, Soren Brage, Tom Yates

## Abstract

**Background:** Although the cardiovascular disease (CVD) benefits of both overall volume and intensity of physical activity (PA) are known, the role of PA intensity, over and above volume, is poorly understood. We aimed to investigate the interplay between PA volume and intensity in relation to incident CVD.

**Methods:** Data were from 88,412 UK Biobank participants without prevalent CVD (58% women) who wore an accelerometer on their dominant wrist for 7 days, from which we estimated total physical activity energy expenditure (PAEE) using population-specific validation. Cox proportional hazards regressions modelled associations between PAEE (kJ/kg/day)] and PA intensity [%MVPA; the fraction of PAEE accumulated from moderate-to-vigorous-intensity PA] with incident CVD, adjusted for potential confounders.

**Results:** There were 4,068 CVD events during 584,568 person-years of follow-up (median 6.8 years). Higher PAEE and higher %MVPA (adjusted for PAEE) were associated with lower rates of incident CVD. In interaction analyses, CVD rates were 17% (95%CI: 8-26%) lower when MVPA accounted for 20% rather than 10% of 15 kJ/kg/d PAEE; equivalent to the difference between a 12-min stroll into a brisk 7-min walk. CVD rates did not differ significantly between values of PAEE when the %MVPA was fixed at 10%. However, the combination of higher PAEE and %MVPA was associated with lower CVD rates. Rates were 24% (10-35%) lower for 20 kJ/kg/d PAEE with 20% from MVPA, and 49% (23-66%) lower for 30 kJ/kg/d with 40% from MVPA (compared to 15 kJ/kg/d PAEE with 10% MVPA).

**Conclusions:** Reductions in CVD risk may be achievable through higher levels of PA volume and intensity, with the role of moderately intense PA appearing particularly important for future CVD risk. Our findings support multiple approaches or strategies to PA participation, some of which may be more practical or appealing to different individuals.

## INTRODUCTION

Regular physical activity (PA), particularly moderate-to-vigorous intensity physical activity (MVPA), is associated with a myriad of health benefits, including lower risk of cardiovascular disease (CVD), cancer, and all-cause mortality (1-3). However, epidemiological evidence used to inform current PA guidelines has relied mostly on self-reported measures of leisure-time or aerobic MVPA, which comprise only a very small proportion of the day and are prone to recall bias and measurement error (4, 5). In contrast, device-based measures of PA can more accurately capture sporadic activity of different intensities throughout the whole waking day, including light-intensity PA (an important contributor to total PA volume (6)), which could enable more specific and targeted PA recommendations.

Several cohort studies are now starting to report findings on the associations between device-based measures of PA with all-cause mortality (7-11), but few have examined associations with incident CVD (12-15). An important recent development has been the ability to process device-based measures of PA to understand the importance of volume and intensity as determinants of disease risk. Elucidating this relationship can be challenging, since PA volume is the product of intensity and time, thus volume and intensity are intrinsically linked. Nevertheless, they represent separate constructs and when used together, can capture different behavioural profiles (16, 17). For example, a person may achieve the same PA volume via a large amount of light-intensity PA (e.g. “pottering about”), or through short periods of higher intensity PA interspersed with high quantities of sedentary time (e.g. “an exerciser” or “active commuter”).

A recent study highlighted the importance of this volume/intensity interaction for all-cause mortality, where a reduced risk was associated with higher volumes of PA, and with higher proportions of a given volume undertaken at a moderate-to-vigorous intensity (7). However, the interplay between volume and intensity has not been clearly elucidated in terms of risk for CVD, with previous studies only reporting associations related to time spent in MVPA or VPA (13, 15). There are supporting mechanisms suggesting that PA intensity may play a specific role in CVD risk, over and above volume, potentially due to greater stimulation and adaptation of cardiorespiratory-related pathways (18-22). Therefore, the interplay between PA volume and intensity warrants further investigation in association with CVD outcomes. Here, we investigate how device-based estimates of PA volume and different PA intensity profiles are associated with incident CVD in UK Biobank, the largest study of accelerometer-measured PA to date.

## METHODS

### Data source and study population

We used data from UK Biobank (application #33266), a population-based prospective cohort study of over 500,000 adults aged 40-69 years, recruited between 2006 to 2010 from across the UK. Methods have been described in detail previously (23). In brief, a sub-sample of 103,686 participants responded to an email for the accelerometer sub-study between June 2013 and December 2015, with PA measurement a median of 5.3 years after their recruitment into the main study (24). The UK Biobank study received ethical approval from the Northwest England Research Ethics Committee (reference 16/NW/0274). Participants gave informed consent before participation.

### Physical activity volume and intensity derived from wrist acceleration

Accelerometry subsample participants were asked to wear a triaxial accelerometer (AX3, Axivity, UK) on their dominant wrist continuously (24 h/day) for seven consecutive days. Measured acceleration was calibrated to local gravity (25) and the movement-related acceleration component isolated from noise and gravity and expressed as the Euclidian Norm Minus One (ENMO) metric (26). Non-wear was quantified as time periods of ≥60 min where the standard deviation of acceleration in each of the three axes was <13 m*g*, which was taken into consideration to minimise diurnal bias when summarising the 5-s epoch time-series to average movement volume and distribution of intensity (25, 26). The average ENMO over 5-s epochs (the intensity time-series) was summarized into average proportions of daily time spent at different movement intensity levels (24). We estimated instantaneous PAEE from wrist movement intensity (27), the time integral of which constitutes total volume of activity as PAEE, as validated against the gold-standard criterion of doubly-labelled water (28) (Table S1). Participants were excluded if their accelerometer record failed calibration (including those not calibrated on their own data), had <3 days of valid wear (defined as >16 h/day), or wear data were not present for each 15-min period of the 24-h cycle (Figure S1). We focussed on two key metrics (Table S1) to summarize total PA volume and intensity, respectively:

- Average daily PAEE (kJ/kg/day) – calculated as the sum of activity-based energy expenditure from all intensity levels.
- Fraction of PAEE from MVPA (%MVPA) – calculated as the sum of energy expenditure from any activity above 125 mg (equivalent to 3 METs) divided by total PAEE.

### Covariate measurement

All participants completed a touchscreen questionnaire and anthropometric assessment at recruitment into the main study, and some participants took part in up to two further touchscreen interviews. Since the accelerometry time-point was used as the analytical baseline for this study, covariate data from the interview undertaken closest to the accelerometry were used (7). Exceptions were: sex and Townsend Index of deprivation (based on postcode) that were only obtained at baseline; ethnicity (assumed not to have changed); and family medical history where a condition was counted if it was reported at any measurement point.

Covariates for this analysis included demographic and lifestyle related characteristics of age, sex, ethnicity (white/non-white), Townsend Index of deprivation, highest educational level achieved (degree or above/any other qualification/no qualification), employment status (unemployed/in paid or self-employment), parental history of CVD or cancer, season of accelerometry wear (using two orthogonal sine functions), alcohol drinking status (never/previous/current), salt added to food (never/sometimes), oily fish intake (never/sometimes), fruit and vegetable intake (a score from 0-4 taking into account questions on cooked and raw vegetables, fresh and dried fruit consumption), processed and red meat intake (average weekly frequency in days per week), and sleep duration (<7, 7-8, >8 h), and a diagnosis of cancer prior to baseline. Prevalent CVD and cancer variables were derived from the self-reported history of heart attack, angina, stroke, or cancer variables, and from hospital episode data (corresponding ICD-10 codes for CVD or cancer I20-25, I60-69, or C00-99; and ICD-9 codes 410-414, 430-439, or 140-199, 201-208, 209.1-209.3, 209.7-209.9). Health-related covariates included blood pressure and cholesterol medications, an insulin prescription or a self-report of doctor diagnosed diabetes, mobility limitations (self-reported longstanding illness or disability or chest pain at rest), and body mass index (BMI) in three categories (<25, 25-30, ≥30 kg/m^2^).

### Ascertainment of incident CVD

Incident non-fatal/fatal CVD was defined as the first appearance of ischaemic heart disease (ICD-10/9 codes I20-25/410-414) or cerebrovascular disease (ICD-10/9 codes I60-69/430-438.9), identified from linkages to Hospital Episode Statistics (HES) or the national death index. Participants who did not experience a cardiovascular disease outcome were censored at death or the end of the study period, as appropriate (England 30/09/2021; Wales 28/02/2018; Scotland 31/07/2021).

### Statistical analyses

All analyses were conducted using Stata v15.1 (StataCorp, TX, USA) and statistical significance was set at *p*<0.05 (two-tailed); results are reported with 95% confidence interval (CI). Participants with CVD prior to accelerometer wear were excluded. We also excluded those who had a CVD event (n=564) within the first year of follow-up, to reduce the risk of reverse causality bias. Using Cox proportional hazard regression models, we first investigated the associations of PAEE and fraction of PAEE from MVPA (the latter adjusted for PAEE) with incident CVD. These models used age as the underlying timescale, and modelled exposures using cubic splines with three evenly-spaced knots. Exposure reference values were chosen as the nearest 5 kJ/kg/day or 5% to the first percentile of the distribution among those who had a CVD event.

Directed acyclic graphs were used to visualise causal assumptions and guide which covariates to include in analyses, and at which level, *a priori* (29). We divided the covariates into two groups based on their likelihood of being on the causal pathway between PA and incident CVD (see Figure S2). As per STROBE recommendations, Model 0 adjusted for sex, with age as the underlying time scale. Model 1 was the main confounder-adjusted model and further included demographic and lifestyle covariates (sex, ethnicity, education level, employment status, Townsend index of deprivation, season of accelerometer wear, dietary variables, alcohol intake, smoking status, average sleep duration, and parental history of cardiovascular disease or cancer) and prevalent cancer. Model 2 additionally adjusted for health-related variables thought to be on the causal pathway, including blood pressure or cholesterol medication use, diabetes diagnosis or insulin prescription, body mass index, and mobility limitation. We used multiple imputation by chained equations (5 imputed datasets) for individuals with missing covariates. All covariates were included in the imputation model as well as the Nelson-Aalen estimate of cumulative baseline hazard of CVD and the incident CVD variable (30).

Interactions between PA volume and intensity were investigated by fitting a spline regression for PAEE and log-transformed %PAEE from MVPA, including interaction terms between the four orthogonal spline variables and %PAEE from MVPA. Using the coefficients, we plotted the fitted spline functions showing the association between PAEE and CVD risk for incremental fractions of PAEE from MVPA (10, 20, 30 and 40%). A 15 kJ/kg/day and 10% PAEE from MVPA reference was chosen for these models. Due to known differences in activity levels by sex in this cohort (24), interaction analyses were also sex-stratified to investigate integrated volume/intensity associations for women and men separately.

#### Sensitivity analyses

Several additional sensitivity analyses were performed, adjusting for covariates in Model 1. To further investigate potential reverse causality bias, we excluded those who had a CVD event/death within 2 years of follow-up or with prevalent cancer at baseline. We also investigated whether results differed when performing complete-case analysis (i.e. without imputation of missing covariate data). Finally, to assess whether the derived measures of PAEE and %PAEE from MVPA used in this analysis provided a similar dose-response association with CVD incidence as more direct measures of PA using acceleration only, we repeated analyses using alternative exposure definitions of PA volume (average ENMO in m*g*) and intensity (intensity gradient; a unitless integrated measure which describes the negative curvilinear relationship between PA intensity and the time accumulated at that intensity (17)). As mentioned, Table S1 provides an overview and more detailed description of all the PA metrics used and the methods to calculate them. The relationships between the different PA volume and intensity metrics are also displayed in Figure S3.

## RESULTS

### Descriptive characteristics

Descriptive characteristics of the 88,412 participants at baseline are shown in Table 1 by incident CVD event and sex. Mean age was 62 (SD, 8) years; mean BMI was 26.6 (SD, 4.5) kg/m^2^; and 58% were women. The age range was similar across sexes, but a higher proportion of women had a BMI in the normal range, had never smoked, took medications, or reported markers of poor health. Activity profiles between sexes were similar on average, but men had slightly lower overall PA volume and spent more time in higher intensity activities. During a median of 6.8 (IQR: 6.2-7.3) years (584,568 person-years) of follow-up, 4,068 CVD events occurred.

**Table 1.**
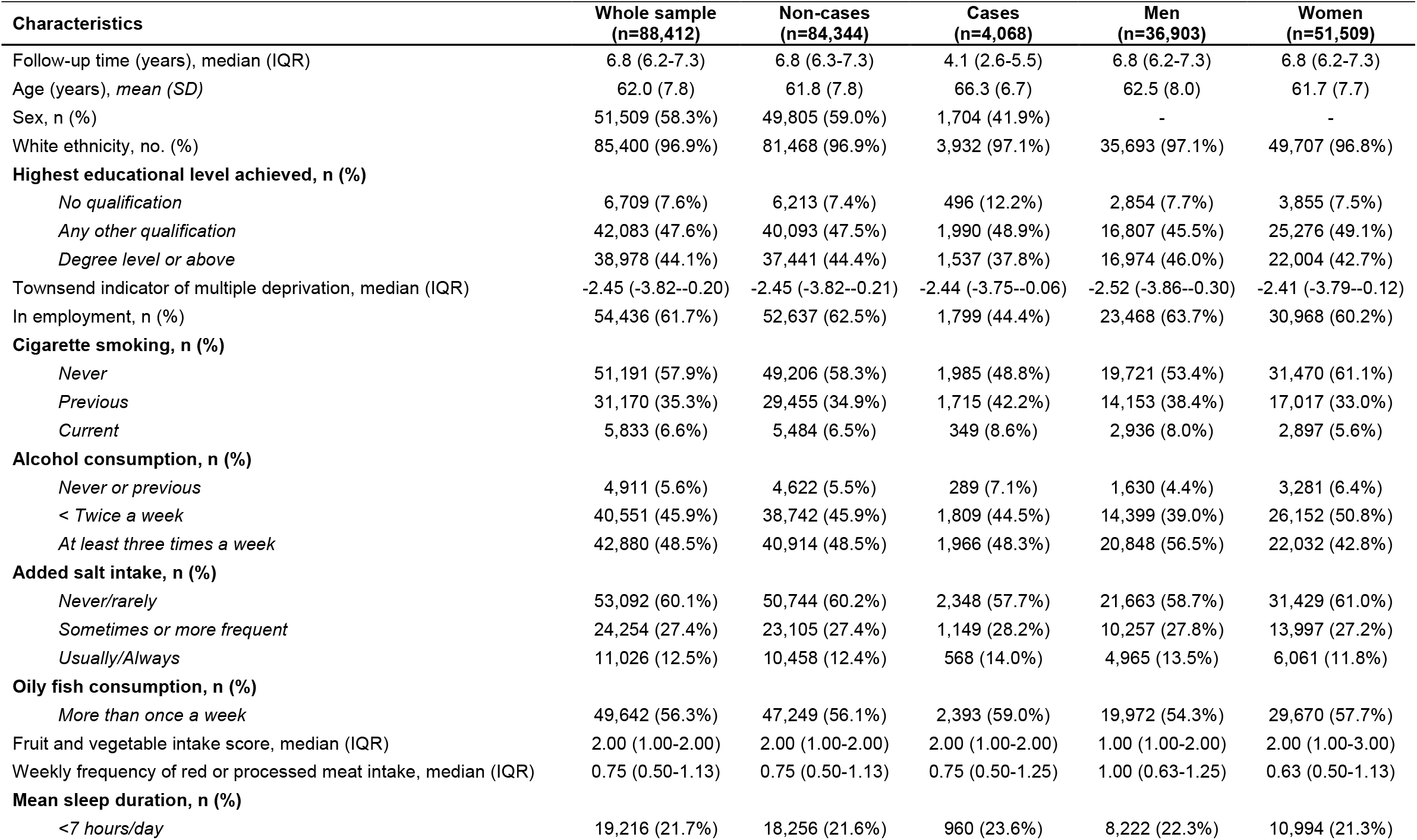
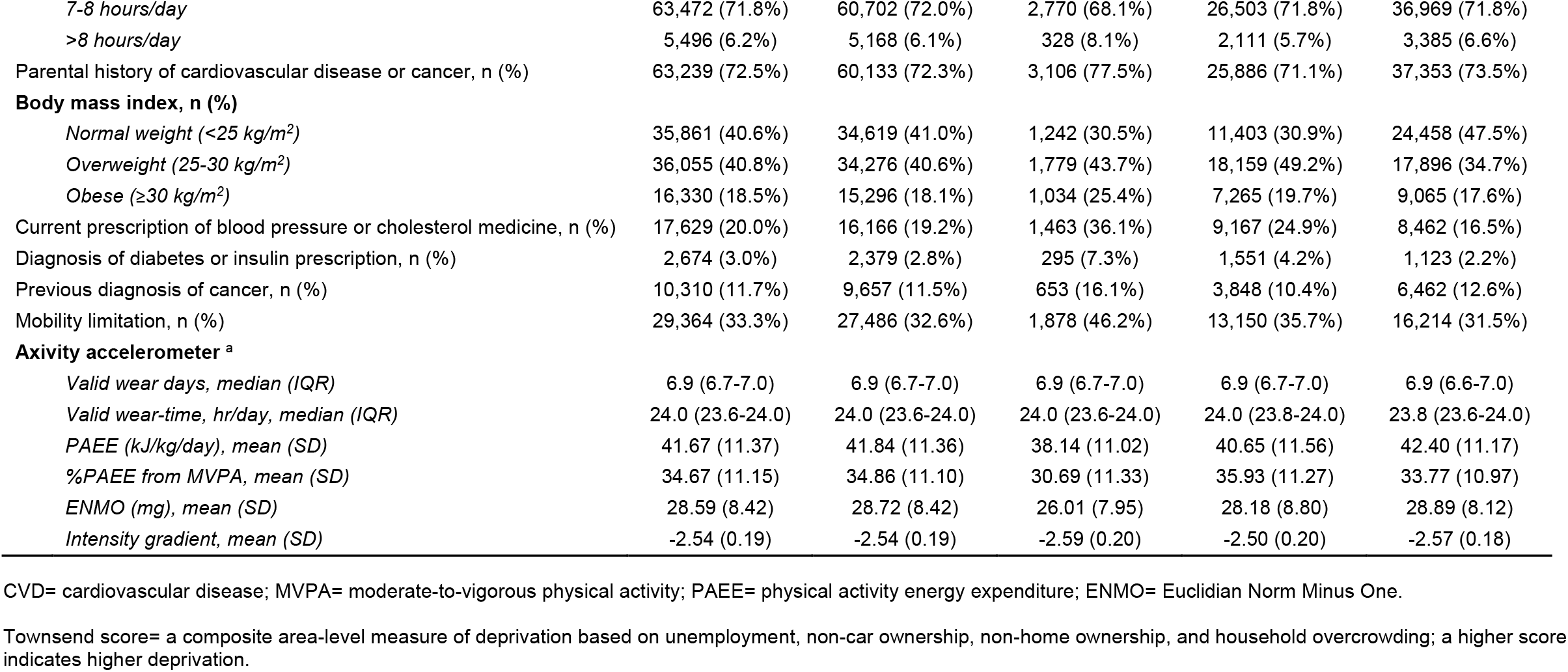
Descriptive characteristics of the whole sample at baseline, by incident CVD status, and by sex.

### Associations of PA volume and intensity

Adjusted for potential confounders and prevalent cancer (model 1), both higher PAEE and %PAEE from MVPA (adjusted for PAEE) were inversely associated with rates of incident CVD (Figure 1; Table 2). Compared to 15 kJ/kg/d, a PAEE of 20 kJ/kg/d was associated with 16% (95%CI: 8-23%) lower rates. PAEE values of 30, 40, and 50 kJ/kg/d were associated with 35% (21-46%), 41% (30-51%), and 47% (37-56%) lower rates, respectively. Compared to accruing 10% of PAEE from MVPA, accruing 20% was associated with 26% (17-35%) lower rates. Accruing 30%, 40%, and 50% of PAEE from MVPA were associated with 40% (31-48%), 48% (39-56%), and 53% (43-61%) lower rates, respectively. Additional adjustment for covariates potentially on the causal pathway (model 2) attenuated all associations slightly.

**Table 2.** Adjusted hazard ratios for incident CVD by volume of PAEE and different fractions of PAEE from MVPA.

| PAEE (kJ/kg/day) | Incident CVD<br>(n=88,412; no. of events=4,068; person years=584,568) |  |  |  |  |  |
| --- | --- | --- | --- | --- | --- | --- |
|  | 15 | 20 | 30 | 40 | 50 | 60 |
| *Model 0* | 1 | 0.80 (0.73-0.87) | 0.57 (0.48-0.69) | 0.50 (0.42-0.59) | 0.44 (0.37-0.53) | 0.41 (0.33-0.49) |
| *Model 1* | 1 | 0.84 (0.77-0.92) | 0.65 (0.54-0.79) | 0.59 (0.49-0.70) | 0.53 (0.44-0.63) | 0.49 (0.40-0.59) |
| *Model 2* | 1 | 0.88 (0.80-0.96) | 0.74 (0.61-0.89) | 0.71 (0.60-0.85) | 0.67 (0.56-0.81) | 0.65 (0.52-0.80) |
| *Model 1 excluding CVD event/death <2yr or prevalent cancer* | 1 | 0.82 (0.74-0.91) | 0.62 (0.49-0.77) | 0.57 (0.46-0.70) | 0.54 (0.43-0.67) | 0.50 (0.39-0.64) |
| *Model 1 complete-case analysis* | 1 | 0.85 (0.78-0.93) | 0.67 (0.55-0.81) | 0.59 (0.50-0.71) | 0.54 (0.45-0.65) | 0.50 (0.41-0.61) |
| **%PAEE from MVPA\*** | 10 | 20 | 30 | 40 | 50 | 60 |
| *Model 0* | 1 | 0.71 (0.63-0.79) | 0.56 (0.49-0.63) | 0.47 (0.42-0.53) | 0.42 (0.36-0.48) | 0.39 (0.31-0.48) |
| *Model 1* | 1 | 0.74 (0.65-0.83) | 0.60 (0.52-0.69) | 0.52 (0.44-0.61) | 0.47 (0.39-0.57) | 0.44 (0.34-0.58) |
| *Model 2* | 1 | 0.77 (0.68-0.87) | 0.66 (0.57-0.77) | 0.60 (0.51-0.71) | 0.56 (0.46-0.68) | 0.54 (0.41-0.71) |
| *Model 1 excluding CVD event/death <2yr or prevalent cancer* | 1 | 0.78 (0.68-0.90) | 0.64 (0.54-0.77) | 0.59 (0.49-0.72) | 0.56 (0.44-0.71) | 0.50 (0.36-0.68) |
| *Model 1 complete-case analysis* | 1 | 0.74 (0.65-0.83) | 0.60 (0.52-0.70) | 0.53 (0.45-0.62) | 0.48 (0.39-0.58) | 0.45 (0.34-0.59) |

Note: Model 1b (n=77,606, no. of events=2,919); Model 1c (n=85,451, no. of events=3,891).
\* %PAEE from MVPA models are additionally adjusted for PAEE. Models 1 and 2 are displayed on Figure 1.
- Model 0 is adjusted for sex (with age as the underlying time scale) only.

**Figure 1.**
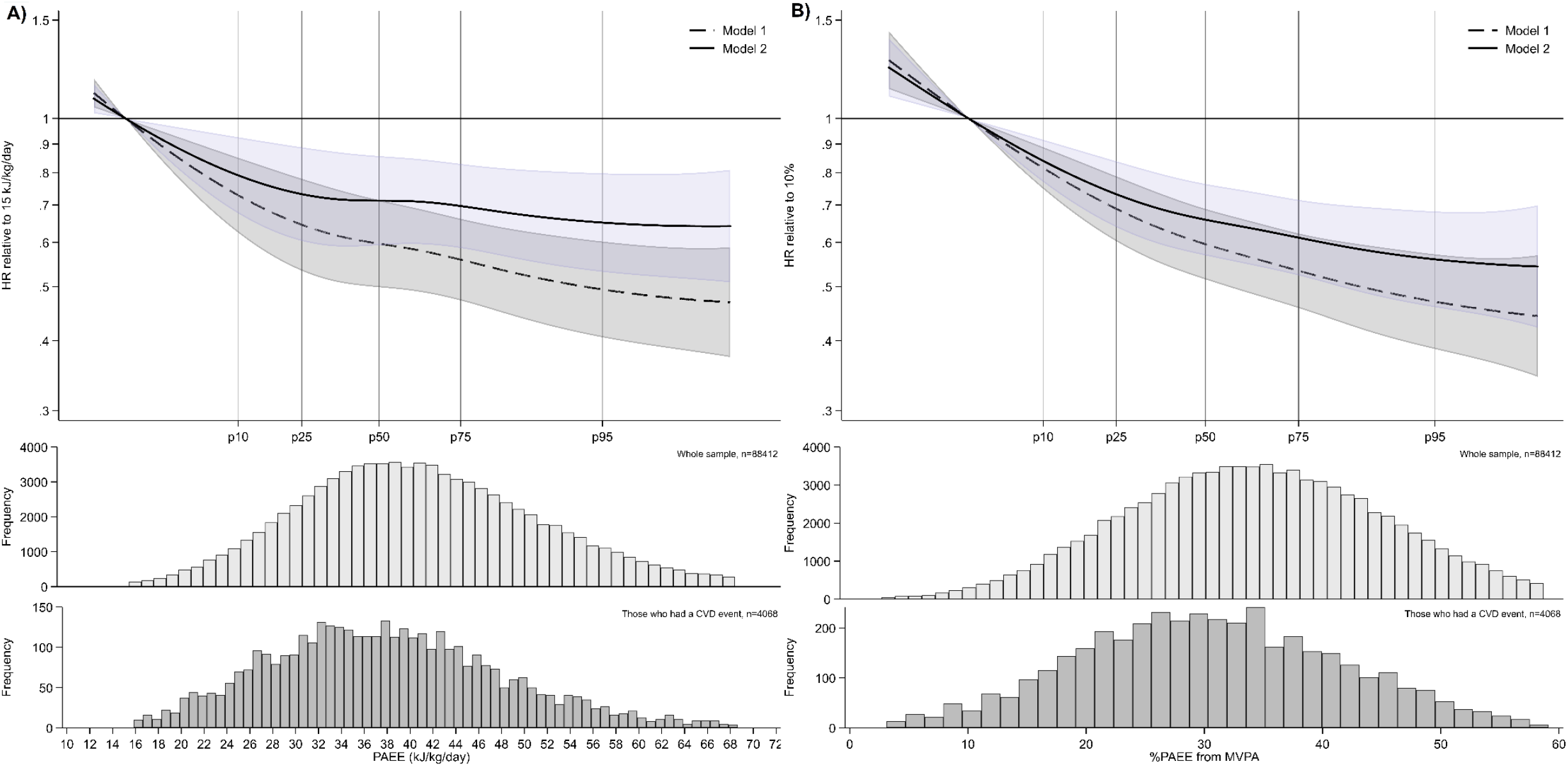
Baseline exposure distribution and adjusted hazard ratios of incident CVD comparing (A) different volumes of PAEE and (B) different fractions of PAEE from MVPA*. * %PAEE from MVPA models (panel B) are additionally adjusted for PAEE - Models were fitted using cubic splines (3 evenly-spaced knots). Adjusted hazard ratios and histogram data shown for values between the 1^st^ or 99^th^ percentiles of the exposure distribution among those who had a CVD event. - Model 1 is adjusted for sex (with age as the underlying time scale), ethnicity, education level, employment status, Townsend index of deprivation, season of accelerometer wear, dietary variables, alcohol intake, smoking status, average sleep duration, parental history of cardiovascular disease or cancer), and prevalent cancer. - Model 2 adjusts for covariates in model 1, with additional adjustment for health-related variables thought to be on the causal pathway (blood pressure or cholesterol medication use, insulin prescription or diagnosed diabetes, body mass index, and mobility limitation). - Further sensitivity analyses are detailed in Table 2 and Figure S5.

### Interaction between PA volume and intensity

In joint volume-intensity analyses, CVD rates were 17% (8-26%) lower when MVPA accounted for 20% rather than 10% of a fixed volume level of 15 kJ/kg/d PAEE (Figure 2; Table 3). CVD rates did not differ significantly with higher values of PAEE when the %PAEE from MVPA was fixed; however, the combination of higher PAEE and %PAEE from MVPA was associated with lower CVD rates. For example, rates were 24% (10-35%) lower for 20 kJ/kg/d PAEE with 20% from MVPA, 29% (8-45%) lower for 30 kJ/kg/d PAEE with 20% from MVPA, and 49% (23-66%) lower for 30 kJ/kg/d with 40% from MVPA (all compared to 15 kJ/kg/d PAEE with 10% MVPA). There was considerable uncertainty around levels of PAEE beyond 40 kJ/kg/day with a >20% fraction of MVPA. Additional adjustment for covariates thought to be on the causal pathway (model 2) slightly attenuated the associations.

**Table 3.** Adjusted hazard ratios of incident CVD for different values of PAEE and the fraction of PAEE from MVPA.

|  |  | Model 1 | Model 2 | Model 1 excluding<br>CVD event/death<br><2yr or prevalent<br>cancer) | Model 1 complete-case analysis |
| --- | --- | --- | --- | --- | --- |
| **n** |  | 88412 |  | 77606 | 85451 |
| **Person-years** |  | 584568 |  | 516559 | 565068 |
| **CVD events** |  | 4068 |  | 2919 | 3891 |
| **PAEE**<br>**(kJ/kg/day)** | **%PAEE**<br>**from MVPA** |  |  |  |  |
| 15 | 10 | 1 (REF) | 1 (REF) | 1 (REF) | 1 (REF) |
|  | 20 | 0.83 (0.74-0.92) | 0.85 (0.76-0.95) | 0.92 (0.81-1.05) | 0.81 (0.72-0.90) |
|  | 30 | N/A | N/A | N/A | N/A |
|  | 40 | N/A | N/A | N/A | N/A |
| 20 | 10 | 0.97 (0.86-1.10) | 0.98 (0.86-1.11) | 0.93 (0.81-1.08) | 0.98 (0.87-1.12) |
|  | 20 | 0.76 (0.65-0.90) | 0.80 (0.68-0.95) | 0.80 (0.66-0.96) | 0.76 (0.65-0.90) |
|  | 30 | 0.66 (0.53-0.83) | 0.72 (0.57-0.90) | 0.73 (0.56-0.95) | 0.66 (0.52-0.82) |
|  | 40 | N/A | N/A | N/A | N/A |
| 30 | 10 | 0.98 (0.78-1.22) | 0.99 (0.79-1.25) | 0.87 (0.65-1.16) | 0.99 (0.78-1.25) |
|  | 20 | 0.71 (0.55-0.92) | 0.78 (0.60-1.01) | 0.67 (0.49-0.91) | 0.72 (0.55-0.94) |
|  | 30 | 0.59 (0.42-0.82) | 0.67 (0.48-0.95) | 0.57 (0.38-0.86) | 0.60 (0.42-0.85) |
|  | 40 | 0.51 (0.34-0.77) | 0.61 (0.40-0.92) | 0.51 (0.31-0.84) | 0.52 (0.34-0.80) |
| 40 | 10 | 1.06 (0.81-1.40) | 1.09 (0.83-1.45) | 0.93 (0.66-1.30) | 1.05 (0.79-1.39) |
|  | 20 | 0.75 (0.60-0.93) | 0.84 (0.67-1.06) | 0.70 (0.54-0.91) | 0.75 (0.59-0.94) |
|  | 30 | 0.61 (0.44-0.83) | 0.72 (0.52-0.99) | 0.59 (0.41-0.87) | 0.61 (0.44-0.85) |
|  | 40 | 0.52 (0.35-0.79) | 0.65 (0.43-0.97) | 0.53 (0.32-0.86) | 0.53 (0.35-0.81) |
| 50 | 10 | N/A | N/A | N/A | N/A |
|  | 20 | N/A | N/A | N/A | N/A |
|  | 30 | 0.60 (0.44-0.83) | 0.72 (0.52-1.00) | 0.62 (0.42-0.90) | 0.60 (0.43-0.84) |
|  | 40 | 0.52 (0.35-0.77) | 0.65 (0.43-0.97) | 0.54 (0.33-0.87) | 0.53 (0.35-0.79) |
| 60 | 10 | N/A | N/A | N/A | N/A |
|  | 20 | N/A | N/A | N/A | N/A |
|  | 30 | N/A | N/A | N/A | N/A |
|  | 40 | 0.54 (0.37-0.78) | 0.68 (0.46-1.01) | 0.56 (0.36-0.89) | 0.54 (0.36-0.80) |

- N/A indicates the specific combination of exposures not between the 1<sup>st</sup> and 99<sup>th</sup> percentiles of the PAEE distribution among those who had a CVD event for that %PAEE from MVPA.
- All hazard ratios are relative to a PAEE of 15 kJ/kg/day mg and a %PAEE from MVPA of 10%. Models 1 and 2 are displayed on Figure 2.

**Figure 2.**
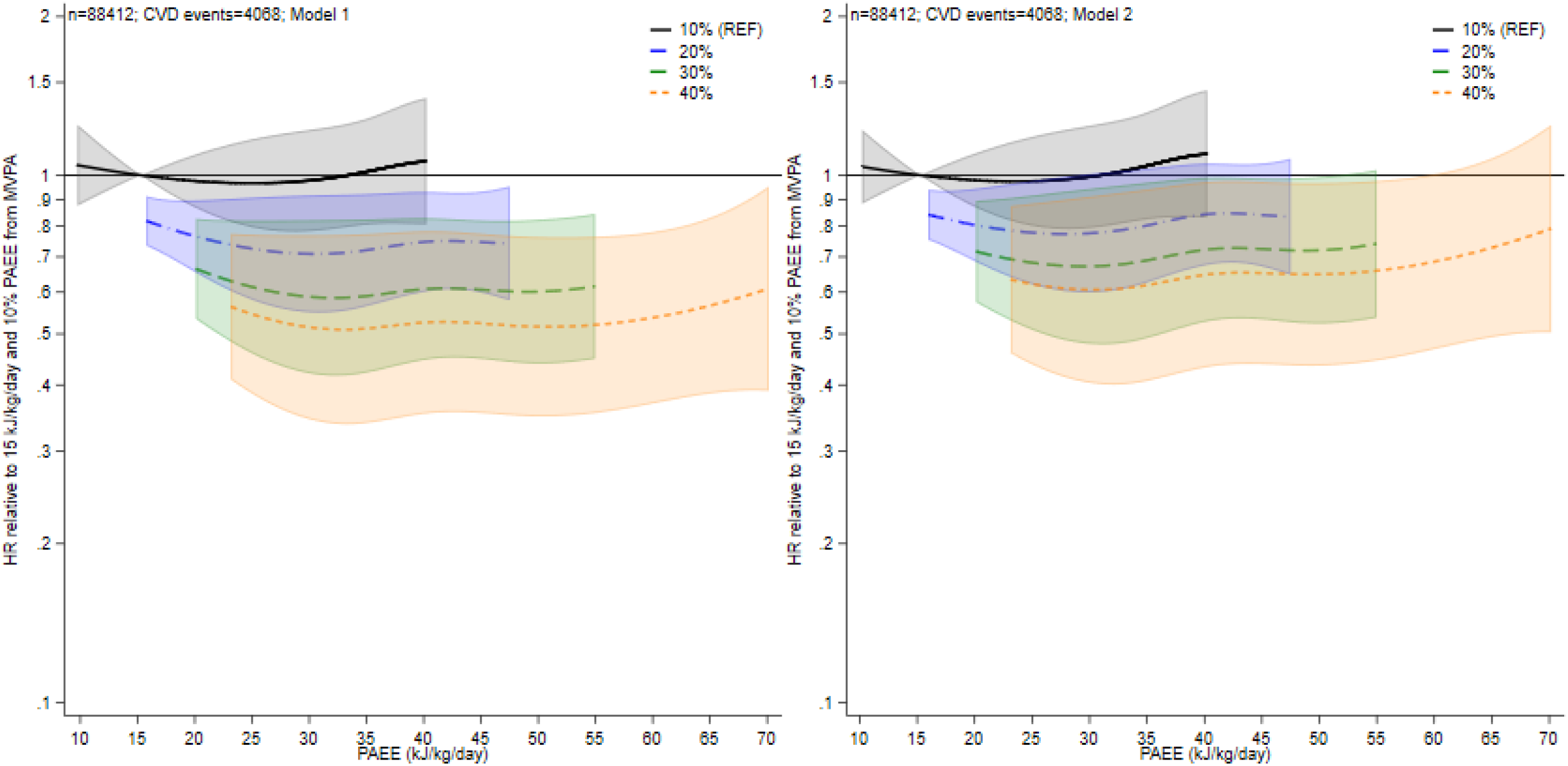
Associations of volume of PAEE and the %PAEE from MVPA with incident CVD. - All hazard ratios are relative to a PAEE of 15 kJ/kg/day and 10% fraction from MVPA (i.e. hazard ratio, 1). - Moving right along each line reflects the hazard ratio for a higher PAEE volume but a constant %PAEE from MVPA. A comparison between lines at a given point on the x-axis therefore reflects the hazard ratio for an increase in intensity but at a constant PAEE. Hazard ratios (95% CI) are shown for values between the 1^st^ or 99^th^ percentiles of the PAEE distribution among those who had a CVD event. - Model 1 is adjusted for sex (with age as the underlying time scale), ethnicity, education level, employment status, Townsend index of deprivation, season of accelerometer wear, dietary variables, alcohol intake, smoking status, average sleep duration, parental history of cardiovascular disease or cancer), and prevalent cancer. - Model 2 adjusts for covariates in model 1, with additional adjustment for health-related variables thought to be on the causal pathway (blood pressure or cholesterol medication use, insulin prescription or diagnosed diabetes, body mass index, and mobility limitation). - Further details are shown in Table 3.

Sex-stratified interaction analyses showed a broadly similar pattern of PAEE and %PAEE from MVPA associations with CVD rates for both men and women (Figure 3, Figure S4 and Table S2), with the lowest rates of CVD seen with higher levels of both PAEE and %PAEE from MVPA.

**Figure 3.**
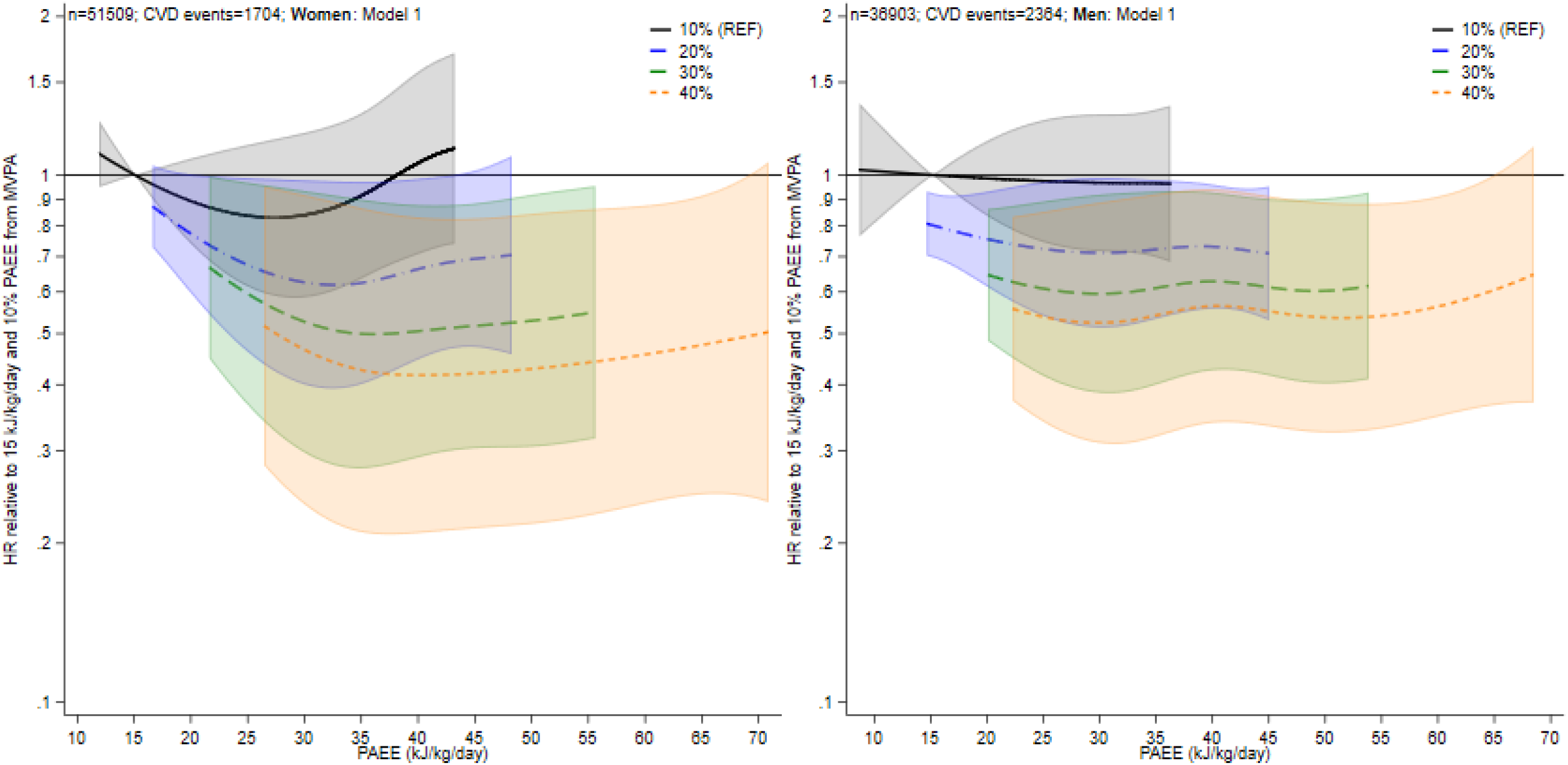
Associations of volume of PAEE the %PAEE from MVPA with incident CVD (model 1), by sex. - All hazard ratios are relative to a PAEE of 15 kJ/kg/day and 10% fraction from MVPA. Moving right along each line reflects the hazard ratio for a higher PAEE volume but a constant %PAEE from MVPA. A comparison between lines at a given point on the x-axis reflects the hazard ratio for an increase in intensity, but a constant PAEE. Hazard ratios shown for values between the 1^st^ or 99^th^ percentiles of the PAEE distribution among those who had a CVD event. - Model 1 is adjusted for sex (with age as the underlying time scale), ethnicity, education level, employment status, Townsend index of deprivation, season of accelerometer wear, dietary variables, alcohol intake, smoking status, average sleep duration, parental history of cardiovascular disease or cancer), and prevalent cancer. - Figure S4 displays results for model 2. Further details are also shown in Table S2.

### Sensitivity analyses

The direction and strength of associations for PAEE and %PAEE from MVPA with CVD rates were consistent when analyses were conducted using acceleration-defined metrics of ENMO and intensity gradient (Figure S5). Excluding participants who had a CVD event within two years of follow-up or with prevalent cancer resulted in similar to slightly attenuated associations (Tables 2 and 3). In addition, results did not materially differ in complete-case analyses.

## DISCUSSION

In this large population-based cohort study of middle-aged adults, we found that a higher volume of PAEE, as well as a higher fraction of PAEE accumulated in MVPA, were both associated with lower rates of incident CVD. The role of activity intensity in future CVD risk was apparent in combined volume-and-intensity interaction analyses. Accumulating 20% rather than 10% of a total PAEE of 15 kJ/kg/d through MVPA was associated with 17% lower CVD rate. This is equivalent to the difference between a 12-min stroll and a brisk 7-min walk; both have the same volume, but the higher intensity of the latter is associated with lower CVD rates. Meanwhile, the role of volume was less clear at fixed intensity fractions, with CVD rates not varying significantly across different activity volumes when the fraction accumulated from MVPA was held constant. The greatest differences in CVD rates were seen when comparing the combination of higher volume and intensity with the lowest levels. For example, accumulating 40% of a total 30 kJ/kg/d PAEE from MVPA was associated with 49% lower CVD rates compared with accumulating 10% of a total 15 kJ/kg/d PAEE from MVPA. Although consistent with the latest PA guidelines and supportive of messages that “every move counts” for improving health outcomes (1, 2), these findings provide additional evidence that intensity may play an important role in minimising CVD risk, over and above total PA volume.

Our results extend upon previous findings using self-reported (8, 31-35) and accelerometer derived (7, 8, 10, 13, 36) measures of PA by examining in more detail the interplay between PA volume and intensity. Using simple, continuous accelerometer-derived metrics of total PAEE and fraction of PAEE from MVPA, we provide a more detailed and integrated perspective on associations with CVD risk, which were previously ambiguous concerning the interactive role of intensity over and above PA volume (13). A notable observation was that when exposures were combined in interaction analyses, the association of PAEE and CVD risk at a given value of %PAEE from MVPA was weaker than when PAEE was the only exposure. Comparing these results with those from similar analyses for all-cause mortality (7), this finding suggests that intensity is relatively more important in minimising CVD risk.

We had anticipated stronger evidence of an association with PA intensity for incident CVD; since higher intensities should theoretically provide greater stimuli (e.g. overload, specificity, and/or relative intensity) for physiological adaptation in functions recognized to specifically influence and maintain cardiorespiratory fitness and vascular function (18-21, 37). This is consistent with previous research showing that self-reported walking pace, a measure of habitual movement intensity and function, is a stronger predictor of CVD mortality than other PA or lifestyle-related factors (38, 39). In addition, it has previously been noted that cardiorespiratory fitness is a cardiovascular vital sign (40), which has been shown to respond particularly to intensity and less so to volume (41). Therefore, it is possible that the relative importance of intensity observed in this study is mediated by improvements in cardiorespiratory fitness and vascular function.

Although it is important to note the inherent inter-relationships between PA volume and intensity (Figure S3), focusing on increasing MVPA and the intensity of habitual movement, such as walking, regardless of the overall daily volume of PA, could have relevance for CVD prevention or targeting for future interventions. Taken together, the public health message is therefore to increase overall volume of activity and, if possible, do so by incorporating more intense activities. Indeed, for any given activity volume (e.g., walking, or the completion of a set list of manual chores), accumulating this volume at higher intensity (e.g., walking faster, or completing chores more quickly/enthusiastically) would also take up less time, which may be particularly attractive for time-poor individuals or for intervention strategies aimed at freeing up time to increase overall PA levels (19).

A key strength of this study is its large sample size, allowing sufficient variation to investigate interactions across the distributions of PA volume and intensity. In addition, the accelerometer-derived metric of PAEE has a strong validation foundation (24, 25) (see Table S1), is easily interpretable, and potentially more applicable to wrist-worn wearable devices for personalised prevention. Although translation of wrist-worn acceleration to energy expenditure does have some limitations, associations with CVD were consistent when analyses were repeated using purely acceleration-based measures of PA volume and intensity (albeit on different exposure scales), providing further confidence in our results. The extensively phenotyped population allowed a comprehensive investigation into possible confounding or mediating influences on the associations between PA volume or intensity with incident CVD; however, residual bias may also have occurred via some unmeasured factors and/or included variables measured with substantial error. We performed several additional sensitivity analyses to investigate and help minimise the potential for reverse causality biases (an important limitation of any observation study) but acknowledge that we cannot fully ameliorate this concern.

Further limitations include the single time-point measure of PA, which limits any potential inferences related to within-person changes or variability in PA over time, and the non-concurrent measurement of covariates and accelerometry. In addition, UK Biobank is not a population-representative cohort (42) and the accelerometer sample may be subject to further selection pressures, which may impact on generalisability. However PA volumes are comparable to national estimates (43) and a previous study showed exposure-outcome associations found in UK Biobank were similar to results in more representative samples (44). Total CVD is a fairly heterogeneous outcome, and we only considered intensity at an absolute level, while intensity relative to maximal capacity may be more critical to driving physiological adaptations (18, 45, 46). Differences in associations for CVD outcomes relative to all-cause mortality (7) could also be related to variations in follow up time and/or greater exclusions for prevalent disease (47), although further sensitivity analyses did not indicate this to be a major factor. Future pooled research should aim to confirm these findings in other populations, consider including repeated accelerometer PA exposures, and incorporate other biomarkers and disease endpoints (including different CVD sub-types or severity) to shed further light on potential mechanisms. Examination of activity volume and intensity interactions in the context of differing levels of adiposity status would also provide valuable insights (48).

## Conclusion

In this large population-based cohort, we show that both higher volumes of PA, and a greater proportion of that volume accumulated as at least moderate intensity, are associated with lower rates of incident CVD in both men and women. The role of activity intensity, over and above that of total volume, also appears to be particularly relevant for CVD risk. These findings align with current PA recommendations and are supportive of messages that “any amount of PA is better than none, and more is better” (i.e., every move counts). They also support multiple approaches to PA participation, some of which may be more practical or appealing to different individuals.

## Data Availability

The UK Biobank resource can be accessed by researchers on application. Variables derived for this study will be returned to the UK Biobank for future applicants to request. No additional data are available.

https://www.ukbiobank.ac.uk/register-apply/

## DECLARATIONS

### Ethics approval and consent to participate

The UK Biobank study received ethical approval from the Northwest England Research Ethics Committee (reference 16/NW/0274). Participants gave informed consent before participation.

### Consent for publication

Not applicable

### Authors’ contributions

PCD, AR, TS, KW, SB, and TY formed the core working group and developed the research question. PCD and TS developed the analysis code, and TS independently replicated the results. PCD ran the final analysis and drafted the manuscript. All authors contributed to the interpretation and revised the manuscript for important intellectual content.

## Acknowledgements

We are grateful to the participants of the UK Biobank Study and those who collected and manage the data.

