## Supplemental material for "Association of Physical Activity Volume and Intensity with Incident Cardiovascular Disease: a UK Biobank Study"

**Supplemental Figure S1:** Flowchart of participant exclusions.

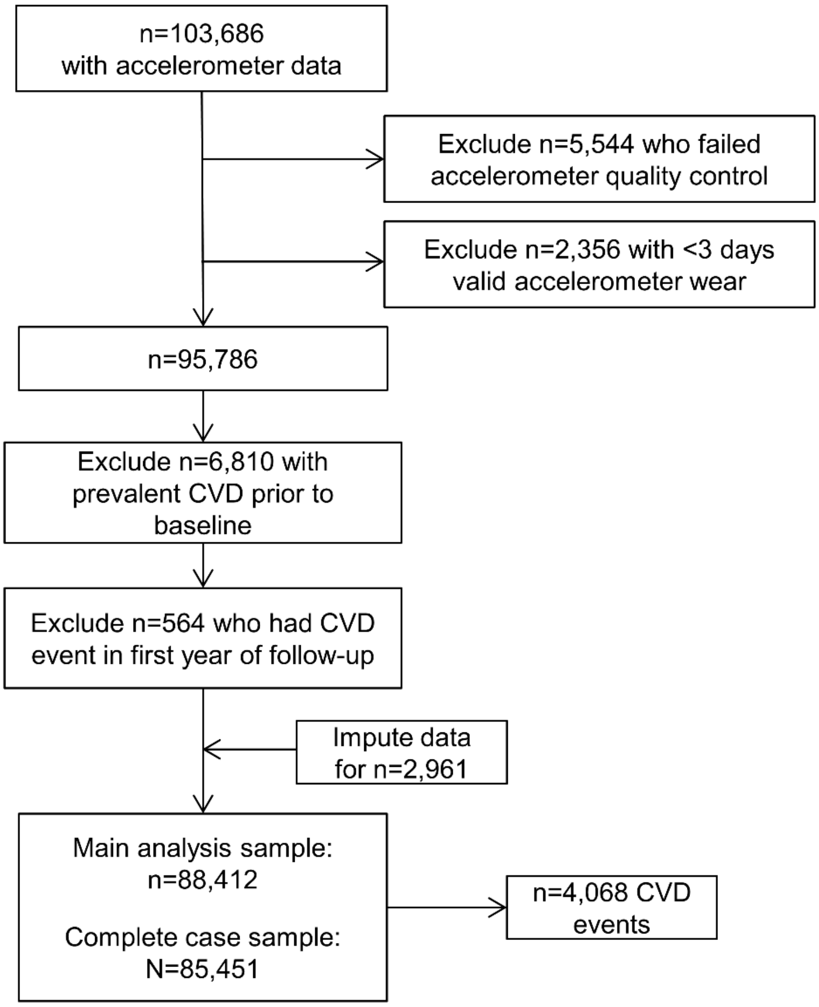

**Supplemental Table S1:** Summary and description of PA volume and intensity variables**.**

| Exposure | Description | Interpretation |
|---|---|---|
| **Volume** | | |
| Physical activity energy expenditure (PAEE), kJ/kg/day | - Time spent in each accelerometer intensity level is converted into a value of PAEE (kJ/kg/day) using equations[1] derived from combined heart rate and trunk acceleration sensors, validated in UK age-matched samples against the gold-standard criterion of doubly labeled water (1, 2).<br>- Predicted PAEE is calculated as the sum of energy expenditure from all intensity levels (3). | - A higher value indicates higher volume of PAEE.<br><br>- note: PAEE and ENMO are very highly correlated metrics of overall PA volume (see *Supplemental Figure S3*) |
| Total PA – Euclidean Norm Minus One (ENMO), m*g* | - Based on the Euclidean Norm (vector magnitude) of the three processed acceleration signals Minus One (with negative values rounded to zero) derived from dominant wrist accelerometry, also referred to as ENMO (4-6).<br>- Summarized as average 24-hour acceleration over all valid days (proxy for total PA). | - A higher value indicates higher total PA (acceleration). |
| **Intensity** | | |
| %PAEE from MVPA | The fraction of PAEE from MVPA (3) is the sum of predicted energy expenditure from wrist-worn accelerometry above 3 METs[2] (threshold of 125 m*g*) divided by total PAEE, expressed as a percentage. | - A higher value indicates higher fraction of PAEE is spent in MVPA. |
| Intensity gradient, unitless | - The intensity gradient describes the negative curvilinear relationship between PA intensity and the time accumulated at that intensity (7) over 24-h.<br>- The intensity gradient is always negative, reflecting the decrease in time accumulated as intensity increases.<br>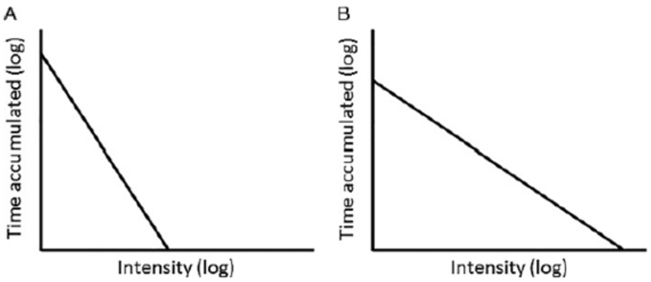<br>**A)** A ***steeper, more negative (lower)*** gradient with a higher constant (y-intercept) showing a steep drop in time accumulated with increasing intensity (left)—a 'poorer' intensity profile.<br><br>**B)** A ***shallower, less negative (higher)*** gradient with a lower constant (y-intercept) showing more time spread across the intensity range (right)—a 'better' intensity profile. | - a higher (less negative) value indicates proportionally more time is habitually spent in higher intensity activities (e.g., brisk walking), or more time spread across the intensity distribution. |

[1] The quadratic equation from White et al. (1) converts dominant wrist accelerometry ENMO processed signal into activity-related energy expenditure measured in J/min/kg: -10.58 + 1.1176*(1.5 + .8517*x) + 2.9418*sqrt((1.5 + .8517*x)) - 0.00059277*((1.5 + .8517*x)^2), where x is the category midpoint in m*g*. This was derived through calibration to PAEE measured by combined heart rate and trunk acceleration in 1695 UK adults (2). This approach has subsequently been validated against total PAEE measured using gold-standard doubly-labelled water in 97 adults (r=0.676) (1).
[2] 1 MET is the standard resting metabolic rate defined as 1.0 kcal/kg/h (8).

**Supplemental Figure S2:** Directed acyclic graphs (DAG) of causal assumptions and potential confounder / adjusted covariates.

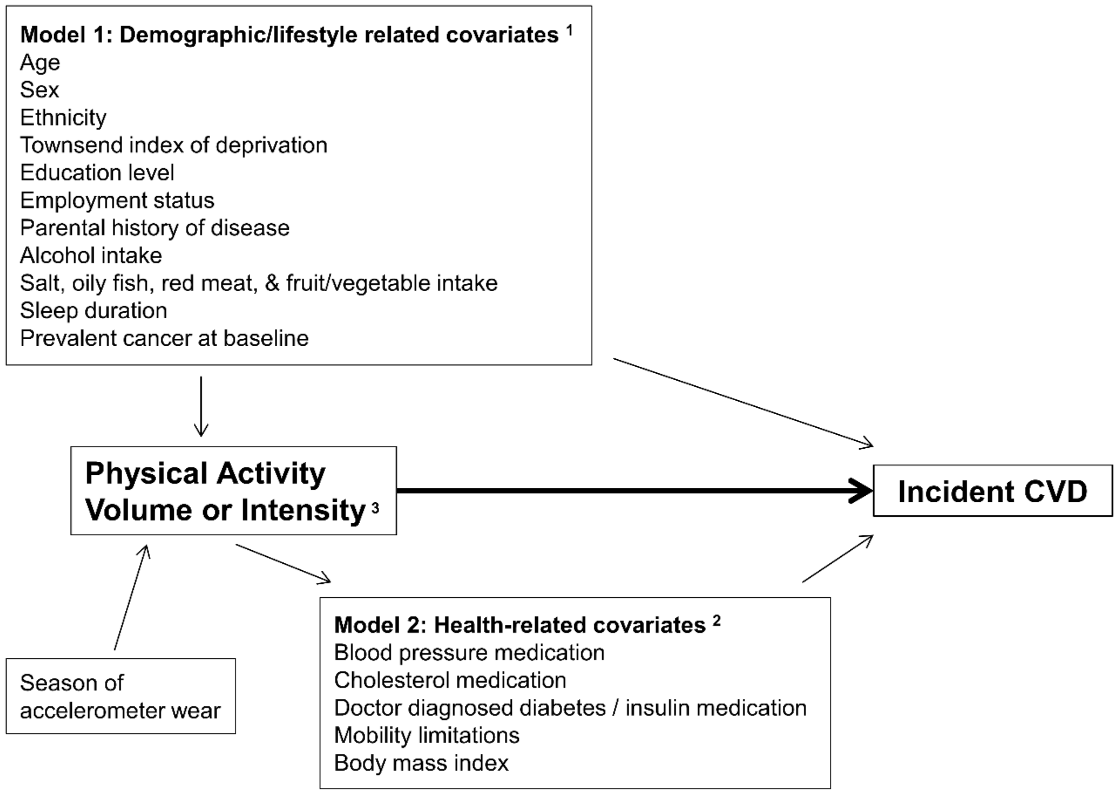


[1] Not on the causal pathway between physical activity and incident CVD.
[2] Potentially on the causal pathway between physical activity and incident CVD.
[3] All %MVPA from PAEE models are adjusted for PAEE.

**Supplemental Figure S3:** Scatter plots showing the relationships between two metrics of PA volume (PAEE and ENMO) vs. intensity (%PAEE from MVPA and intensity gradient), respectively. Correlation coefficients are also displayed between all PA variables.

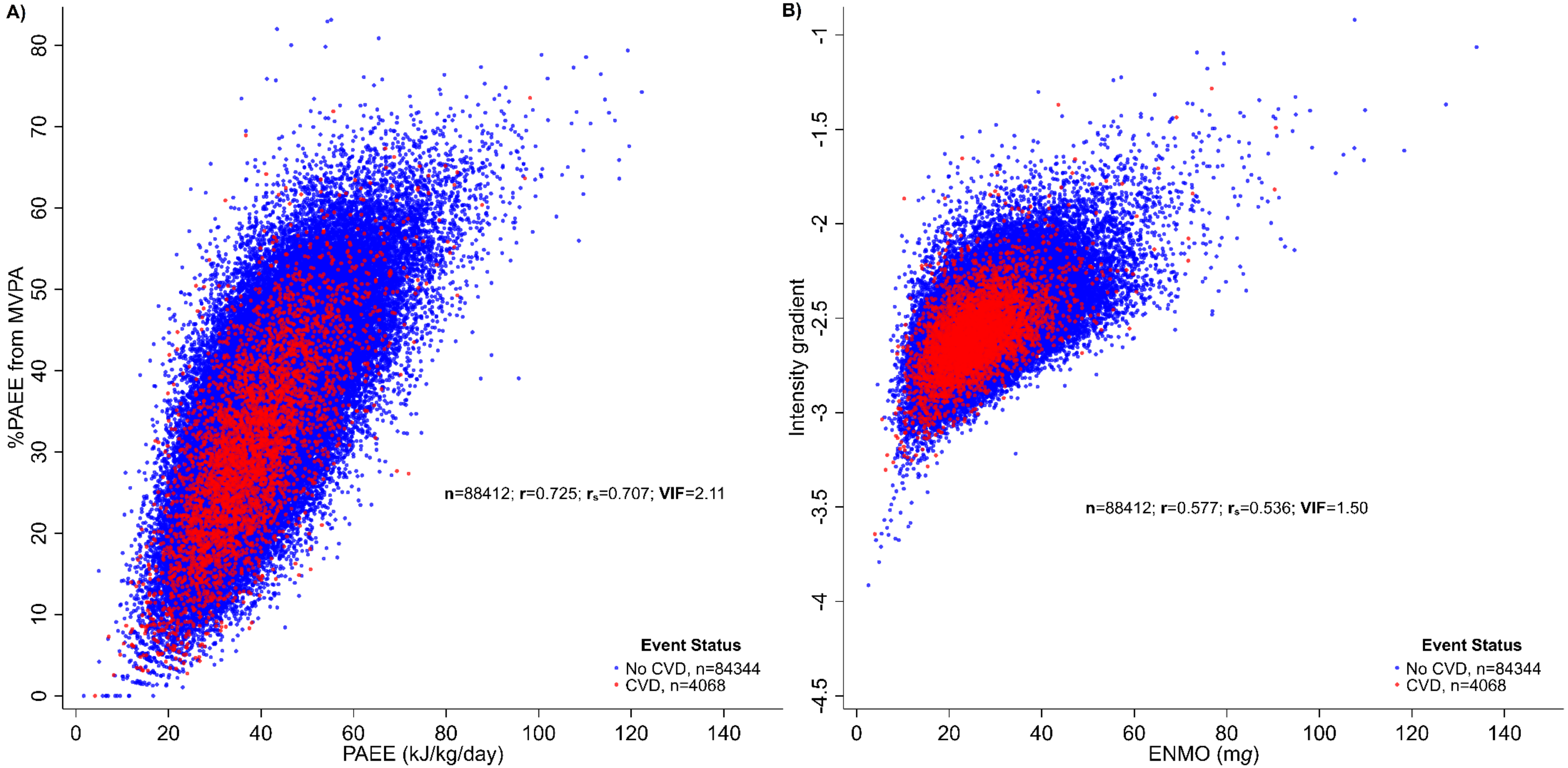


| Pearson's (r) | **ENMO** | **PAEE** | **IG** | **%MVPA from PAEE** |
|---|---|---|---|---|
| **ENMO** | 1 | | | |
| **PAEE** | .966 | 1 | | |
| **Intensity gradient** | .577 | .478 | 1 | |
| **%MVPA from PAEE** | .763 | .725 | .738 | 1 |

| Spearman's ($r_s$) | **ENMO** | **PAEE** | **IG** | **%MVPA from PAEE** |
|---|---|---|---|---|
| **ENMO** | 1 | | | |
| **PAEE** | .978 | 1 | | |
| **Intensity gradient** | .536 | .465 | 1 | |
| **%MVPA from PAEE** | .752 | .707 | .736 | 1 |

r=Pearson's correlation coefficient; $r_s$=Spearman's correlation coefficient; VIF=variance inflation factor; MVPA=moderate-to-vigorous intensity PA; PAEE=physical activity energy expenditure; ENMO=Euclidean Norm Minus One; IG=intensity gradient. See Table S1 for a more detailed description of the PA volume/intensity metrics.

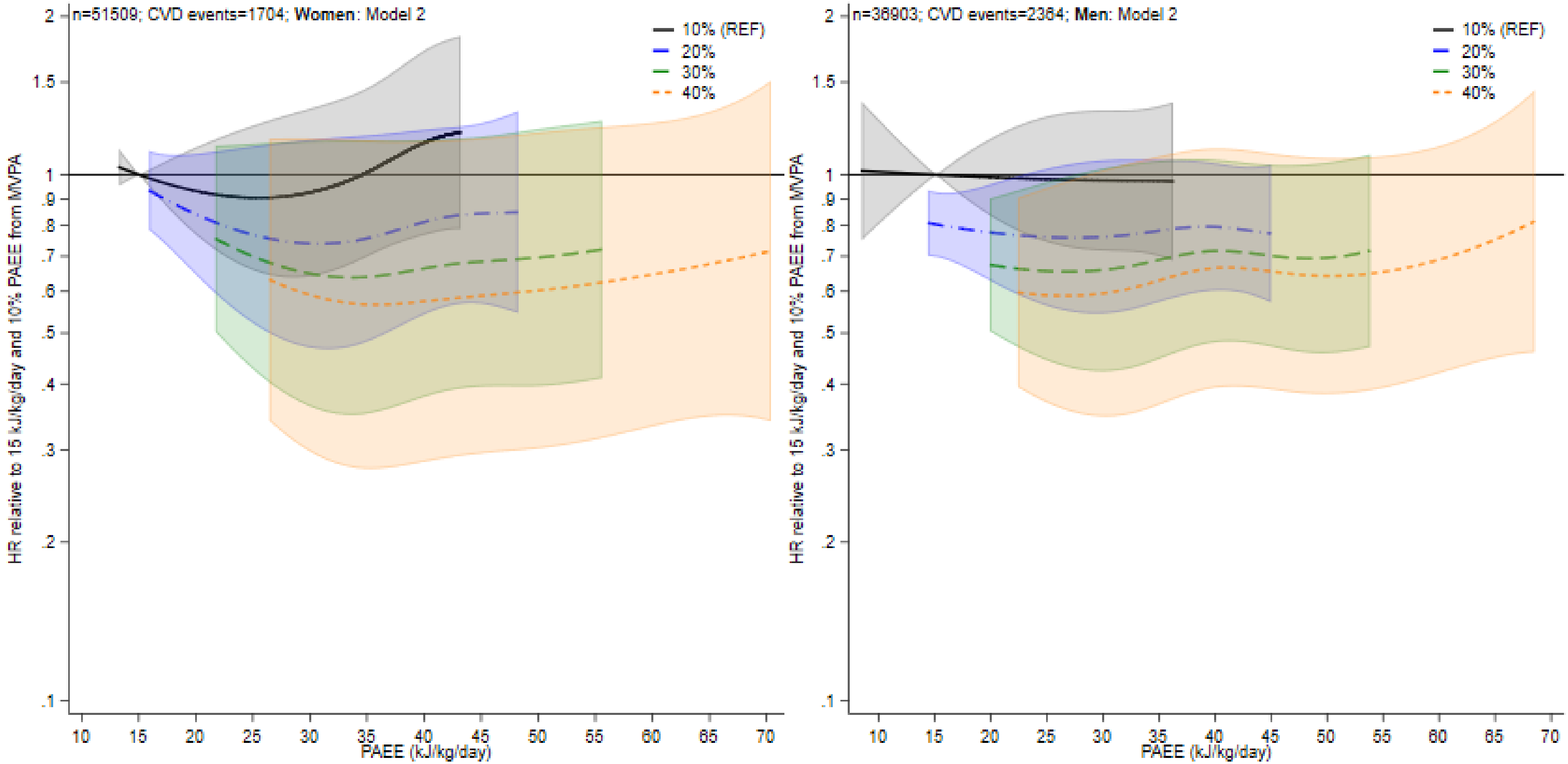


**Supplemental Figure S4.** Associations of volume of PAEE the %PAEE from MVPA with incident CVD (model 2), by sex.

- All hazard ratios are relative to a PAEE of 15 kJ/kg/day and 10% fraction from MVPA (i.e. hazard ratio, 1).
- Moving right along each line reflects the hazard ratio for a higher PAEE volume but a constant %PAEE from MVPA. A comparison between lines at a given point on the x-axis therefore reflects the hazard ratio for an increase in intensity but at a constant PAEE. Hazard ratios (95% CI) are shown for values between the 1st or 99th percentiles of the PAEE distribution among those who had a CVD event.

- Model 2 is adjusted for ethnicity (with age as the underlying time scale), education level, employment status, Townsend index of deprivation, season of accelerometer wear, dietary variables, alcohol intake, smoking status, average sleep duration, and parental history of cardiovascular disease or cancer), prevalent cancer, blood pressure or cholesterol medication use, insulin prescription or diagnosed diabetes, body mass index, and mobility limitation.
- Figure 3 displays results for model 1. Further details are shown in Table S2.

**Supplemental Table S2.** Adjusted hazard ratios of incident CVD for different values of volume of PAEE and the fraction of PAEE from MVPA, stratified by sex.

| | | Women | | Men | |
|---|---|---|---|---|---|
| | | Model 1 | Model 2 | Model 1 | Model 2 |
| **n** | | 51509 | | 36903 | |
| **Person-years** | | 342683 | | 241884 | |
| **CVD events** | | 1704 | | 2364 | |
| **PAEE** | **%PAEE from MVPA** | | | | |
| 15 | 10 | 1 (REF) | 1 (REF) | 1 (REF) | 1 (REF) |
| | 20 | 0.96 (0.81-1.14) | 0.80 (0.70-0.93) | 0.95 (0.81-1.13) | 0.80 (0.70-0.93) |
| | 30 | N/A | N/A | N/A | N/A |
| | 40 | N/A | N/A | N/A | N/A |
| 20 | 10 | 0.90 (0.77-1.07) | 0.99 (0.83-1.16) | 0.93 (0.77-1.13) | 0.99 (0.84-1.17) |
| | 20 | 0.78 (0.62-1.00) | 0.76 (0.61-0.93) | 0.84 (0.65-1.10) | 0.78 (0.63-0.96) |
| | 30 | N/A | N/A | 0.79 (0.56-1.13) | 0.67 (0.50-0.90) |
| | 40 | N/A | N/A | N/A | N/A |
| 30 | 10 | 0.85 (0.60-1.20) | 0.97 (0.71-1.31) | 0.93 (0.64-1.33) | 0.98 (0.72-1.33) |
| | 20 | 0.63 (0.41-0.97) | 0.71 (0.51-0.99) | 0.74 (0.47-1.16) | 0.76 (0.54-1.07) |
| | 30 | 0.53 (0.31-0.93) | 0.60 (0.38-0.92) | 0.65 (0.36-1.16) | 0.66 (0.42-1.03) |
| | 40 | 0.47 (0.25-0.91) | 0.52 (0.31-0.89) | 0.59 (0.30-1.17) | 0.60 (0.35-1.02) |
| 40 | 10 | 1.07 (0.72-1.58) | 0.95 (0.64-1.40) | N/A | N/A |
| | 20 | 0.67 (0.46-0.98) | 0.73 (0.56-0.96) | 0.81 (0.54-1.22) | 0.80 (0.60-1.05) |
| | 30 | 0.51 (0.30-0.87) | 0.63 (0.42-0.93) | 0.66 (0.38-1.16) | 0.72 (0.48-1.07) |
| | 40 | 0.42 (0.22-0.83) | 0.56 (0.34-0.94) | 0.58 (0.29-1.16) | 0.66 (0.39-1.12) |
| 50 | 10 | N/A | N/A | N/A | N/A |
| | 20 | N/A | N/A | N/A | N/A |
| | 30 | 0.54 (0.32-0.91) | 0.60 (0.40-0.91) | 0.70 (0.40-1.22) | 0.70 (0.46-1.06) |
| | 40 | 0.43 (0.22-0.84) | 0.54 (0.32-0.89) | 0.60 (0.30-1.20) | 0.64 (0.38-1.08) |
| 60 | 10 | N/A | N/A | N/A | N/A |
| | 20 | N/A | N/A | N/A | N/A |
| | 30 | N/A | N/A | N/A | N/A |
| | 40 | 0.46 (0.25-0.87) | 0.56 (0.35-0.91) | 0.65 (0.33-1.25) | 0.69 (0.42-1.13) |

- N/A indicates the specific combination of exposures not between the 1st and 99th percentiles of the PAEE distribution among those who had a CVD event for that %PAEE from MVPA.
- All hazard ratios are relative to a PAEE of 15 kJ/kg/day mg and a %PAEE from MVPA of 10%. Models 1 and 2 are displayed on Figure 3 and Figure S4.

- Model 1 is adjusted for ethnicity (with age as the underlying time scale), education level, employment status, Townsend index of deprivation, season of accelerometer wear, dietary variables, alcohol intake, smoking status, average sleep duration, parental history of cardiovascular disease or cancer), and prevalent cancer.
- Model 2 adjusts for covariates in model 1, with additional adjustment for health-related variables thought to be on the causal pathway (blood pressure or cholesterol medication use, insulin prescription or diagnosed diabetes, body mass index, and mobility limitation).

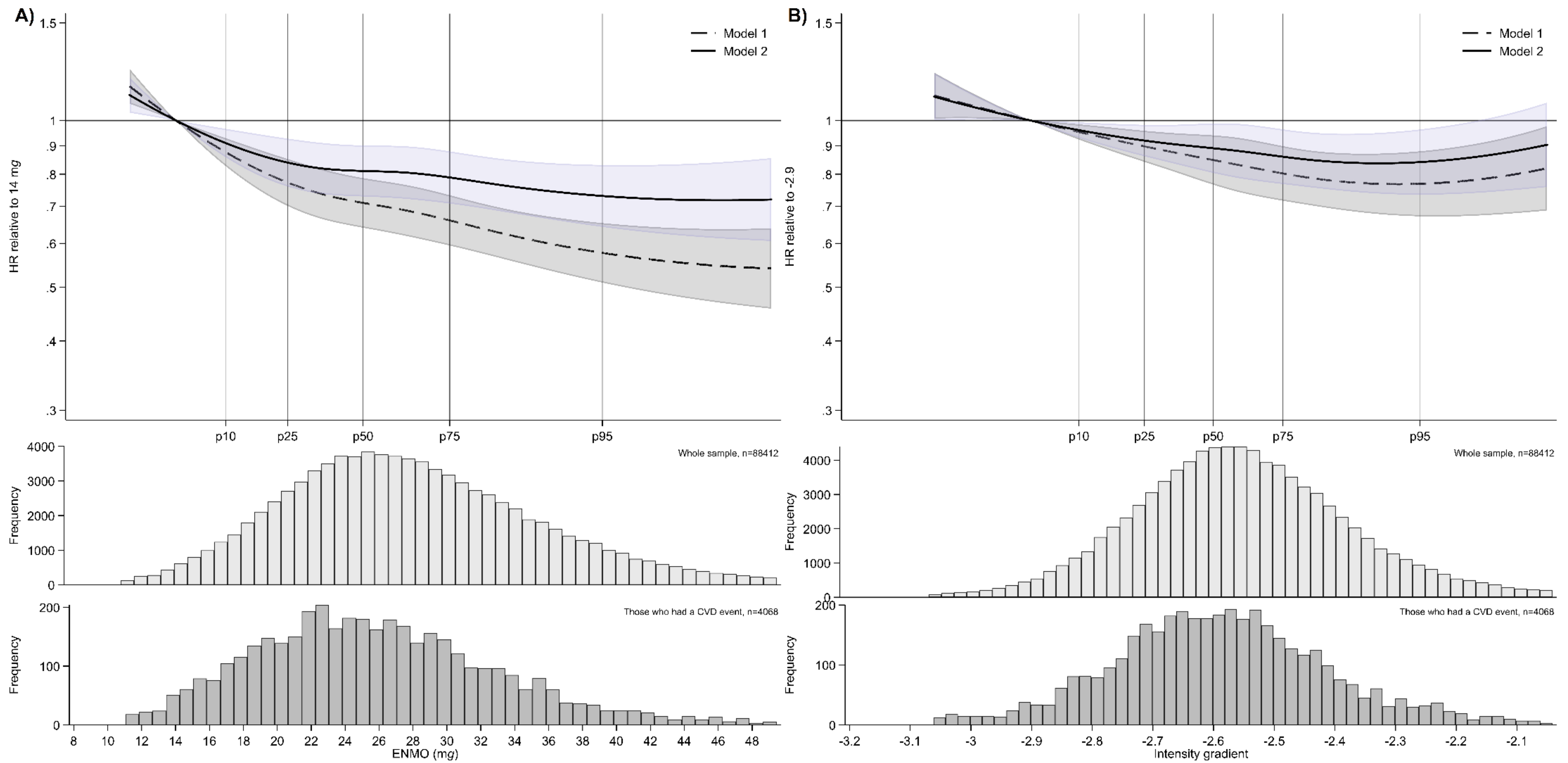


**Supplemental Figure S5.** Baseline exposure distribution and adjusted hazard ratios of incident CVD comparing (A) different amounts of ENMO (a measure of total PA) and (B) different levels of intensity gradient* (a measure of the intensity distribution of PA).

* Intensity gradient models (panel B) are additionally adjusted for total PA – defined here as Euclidean Norm Minus One (ENMO). A higher (less negative) intensity gradient (intensity distribution of PA) indicates more time is habitually spent in higher intensity activities (e.g., brisk walking) over a day.
- Models were fitted with the use of restricted cubic splines (3 evenly-spaced knots). Adjusted hazard ratios and histogram data shown for values between the 1st or 99th percentiles of the exposure distribution among those who had a CVD event.

- Model 1 is adjusted for sex (with age as the underlying time scale), ethnicity, education level, employment status, Townsend index of deprivation, season of accelerometer wear, dietary variables, alcohol intake, smoking status, average sleep duration, parental history of cardiovascular disease or cancer), and prevalent cancer.
- Model 2 adjusts for covariates in model 1, with additional adjustment for health-related variables thought to be on the causal pathway (blood pressure or cholesterol medication use, insulin prescription or diagnosed diabetes, body mass index, and mobility limitation).
- A higher (less negative) intensity gradient value indicates more time is habitually spent in higher intensity activities (e.g., brisk walking).
- See Table S1 for a more detailed description of the PA volume and intensity metrics and the methods used. The relationships between the different PA volume/intensity metrics are also displayed in Figure S3.
